# Orbitofrontal cortex hypergyrification in hallucinating schizophrenia patients: surface ratio as a promising brain biomarker

**DOI:** 10.1101/2024.02.19.24303035

**Authors:** Christian Núñez, Christian Stephan-Otto, Alexandra Roldán, Eva Mª Grasa, Mª José Escartí, Eduardo J. Aguilar García-Iturrospe, Gracián García-Martí, Maria de la Iglesia-Vaya, Juan Nacher, Maria J. Portella, Iluminada Corripio

## Abstract

**Background:** There has been increasing interest in the study of brain gyrification in schizophrenia since it may provide additional useful information on the cytoarchitecture and connectivity of the brain. Various methods have been developed to estimate brain gyrification that, so far, have yielded mixed and inconclusive results in schizophrenia studies. To the best of our knowledge, an alternative method to compute brain gyrification, known as surface ratio (SR), has not yet been applied to a schizophrenia sample. Our aim in this study was to assess whether SR could provide new insights on the brain structure of schizophrenia patients and the severity of symptoms. We also computed a more established brain gyrification measure, namely absolute mean curvature (AMC), for comparison.

**Method:** We processed and analyzed a total of 63 magnetic resonance images, 25 from schizophrenia patients with treatment-resistant auditory verbal hallucinations (SCH-H), 18 from schizophrenia patients without hallucinations (SCH-NH), and 20 from healthy controls (HC). We estimated brain gyrification with SR and AMC employing CAT software.

**Results:** The SR measure mainly revealed that SCH-H patients had a more folded orbitofrontal cortex than SCH-NH patients and HC. Gyrification in this region was also negatively associated with positive symptoms, specifically with the delusions and conceptual disorganization items, only in the SCH-H group. Conversely, SCH-NH and HC showed more SR than SCH-H in other frontal areas. As for the AMC measure, we identified two areas where HC showed more gyrification than SCH-H patients, but no relationships arose with symptoms.

**Discussion:** We hypothesize that the hypergyrification of the orbitofrontal cortex displayed by SCH-H patients, as captured by the SR measure, suggests aberrant and/or excessive wiring in these patients, which in turn could give rise to auditory verbal hallucinations. Alternatively, we comment on potential compensatory mechanisms that may better explain the negative association between orbitofrontal gyrification and positive symptomatology. Importantly, the estimation of brain gyrification with the SR measure seems to capture the most relevant differences and associations, making it a promising biomarker in schizophrenia research.

## Introduction

The study of brain gyrification, or cortical folding, may provide interesting insights on the cytoarchitecture of the brain (Fischl et al. 2008; Ronan and Fletcher 2015), its connectivity (Hogstrom et al. 2013), and its synaptic density (Howes et al. 2023), which could be altered in psychotic disorders. Even though gyrification has not been as widely studied as gray matter volume or cortical thickness, it has recently gained some attention in the study of schizophrenia and related disorders, particularly since popular neuroimaging tools, such as FreeSurfer (https://surfer.nmr.mgh.harvard.edu) and CAT (Gaser et al. 2022; https://neuro-jena.github.io/cat), have automated the computation of this parameter. Methods to measure brain gyrification may be roughly divided into two categories (Rabiei et al. 2017). One of these categories includes those methods based on surface area, such as the local gyrification index proposed by Schaer et al. (2008), which computes the ratio between the outer surface and the pial surface of the brain in a circular region of interest. The second category encompasses methods based on cortical curvature, such as the absolute mean curvature (AMC) method proposed by Luders et al. (2006), which estimates brain gyrification based on curvature measurements throughout the cortical surface.

Some authors have reviewed the available literature on the study of cortical gyrification in schizophrenia disorders (Matsuda and Ohi 2018; Sasabayashi et al. 2021; Zakharova et al. 2021; Howes et al. 2023), which has mostly employed the local gyrification index based on the previously described method of Schaer et al. (2008). The results are mixed, and both hypogyrification and hypergyrification in widespread areas of the brain have been reported. Results derived from studies examining cortical gyrification based on the AMC method are also inconclusive; while some studies have found higher gyrification in patients with schizophrenia in many brain areas, such as the visual cortex (Schultz et al. 2013) and the frontal and temporal lobes (Nenadic et al. 2015; Spalthoff, Gaser, and Nenadić 2018; Otte et al. 2022), regions with lower gyrification have also been reported in these patients (Madeira et al. 2020; Sheffield et al. 2021; Otte et al. 2022). In light of these inconsistencies, it has been hypothesized that younger patients, or those in the earlier stages of the illness, tend to show higher cortical gyrification with respect to healthy controls, while older patients—in their mid-thirties and beyond—show lower cortical gyrification (Sasabayashi et al. 2021; Howes et al. 2023). Other studies have examined the relationship between gyrification and schizophrenia symptoms. While Sasabayashi et al. (2017) reported an association between positive symptoms and hypergyrification of the right temporal pole, insula, and parahippocampal gyrus, Kubera et al. (2018) found that patients with persistent auditory hallucinations presented higher gyrification of the right precuneus and superior parietal cortex, as well as lower gyrification of Broca’s and other frontal areas, compared with patients without auditory hallucinations. On the other hand, hypergyrification of the brain has been found to be associated with less severe negative symptomatology (Takayanagi et al. 2019; Yunzhi et al. 2022; Gao et al. 2023).

Yet another method to estimate brain gyrification, known as surface ratio (SR) or Toro’s gyrification index, was proposed by Toro et al. (2008). Even though the measurements are different, the SR is conceptually similar to the method proposed by Schaer (White et al. 2010; Schaer et al. 2012; Rabiei et al. 2017). The CAT software computes an adaptation of this measure, which is defined as the ratio between the central surface—located between the white matter and the pial surfaces—area contained in a sphere and the area of a disk of the same radius (Gaser et al. 2022). To the best of our knowledge, the SR has not yet been assessed in a sample of patients with a psychotic disorder. However, various studies have reported the utilization of this measure in samples of healthy people (Parker Jones, Alfaro-Almagro, and Jbabdi 2018; Kalantar-Hormozi et al. 2023), healthy people with subclinical psychosis-related symptoms (Dai 2023), autism (Ziolkowski 2023), and attention deficit hyperactivity disorder (Ediriarachchi et al. 2022).

Here we aimed to assess whether SR could provide useful information on brain folding abnormalities not detected by more traditional methods. Hence, we applied the SR measure in a schizophrenia sample that included both patients with treatment-resistant auditory verbal hallucinations (SCH-H) and patients without hallucinations (SCH-NH). Our main objective was to precisely locate the potential brain gyrification differences captured by the SR measure between patients with schizophrenia and healthy controls (HC). Additionally, we also aimed to find potential associations between SR and clinical symptomatology in the group of patients with schizophrenia. Finally, we compared the SR results with those obtained with a more established brain gyrification measure, namely the AMC (Luders et al. 2006). Given that results on brain gyrification in schizophrenia have been rather inconsistent and that this is the first study to use the SR measure in a schizophrenia sample, the analyses were mainly exploratory.

## Methods

### Participants

We analyzed a total of 63 participants (25 SCH-H, 18 SCH-NH, and 20 HC), who were recruited from the Psychiatry Department at Hospital de Sant Pau in Barcelona, Spain. SCH-H and SCH-NH patients met the DSM-IV-TR criteria for schizophrenia. The SCH-H group included patients with treatment-resistant auditory verbal hallucinations—that is, patients with daily presence of auditory verbal hallucinations during the past year despite at least two trials of antipsychotic medication, with different D_2_ binding profiles, at chlorpromazine-equivalent doses of 600 mg/day. The SCH-NH group included patients without a history of hallucinations who showed good response to treatment, and not displaying acute psychotic symptoms in the past 12 months. Exclusion criteria for both groups of patients were the presence of any neurological disorder that could explain the current psychopathology, mental retardation, and substance use (except tobacco, alcohol, and cannabis). HC participants were recruited from the community, and the exclusion criteria were having a history of medical or psychiatric disorders, substance use (except tobacco, alcohol, and cannabis), and a family history of psychiatric disorders.

### Neuroimaging data acquisition and processing

We obtained high-resolution T1-weighted structural images for all the participants, who were scanned in a 3T Philips Achieva scanner employing the following acquisition parameters: TR = 13 ms; TE = 7.4 ms; flip angle = 8°; field of view = 23 cm; acquisition matrix = 256 x 256; slice thickness = 1 mm. All the structural images were acquired on the same day as the clinical visit. Only two patients—one of them belonging to the SCH-H group and the other to the SCH-NH group—had their MRI acquired 1 and 14 days after the clinical visit, respectively. We processed and analyzed all the images with CAT (v12.7, r1742) (Gaser et al. 2022), a toolbox for SPM12 (https://www.fil.ion.ucl.ac.uk/spm/software/spm12/), running under MATLAB (Release 2020b, The MathWorks, Inc., Natick, MA, USA). Image processing included the segmentation of the gray matter, white matter, and cerebrospinal fluid tissues, as well as the reconstruction of the cortical surface. After image processing, we generated two different kinds of brain gyrification maps for each participant: SR maps and AMC maps, based on the methods of Toro et al. (2008) and Luders et al. (2006), respectively. We then resampled these brain gyrification maps to a high-resolution 164k mesh, and we smoothed them with a 20 mm full width at half maximum (FWHM) filter. We used these resampled and smoothed brain gyrification maps in the surface-based morphometry (SBM) analyses explained in the *Statistical analysis* section. We employed the Desikan-Killiany (DK40) atlas (Desikan et al. 2006) to precisely locate our results. Figure 1 depicts how SR and AMC are calculated.

**Figure 1.**
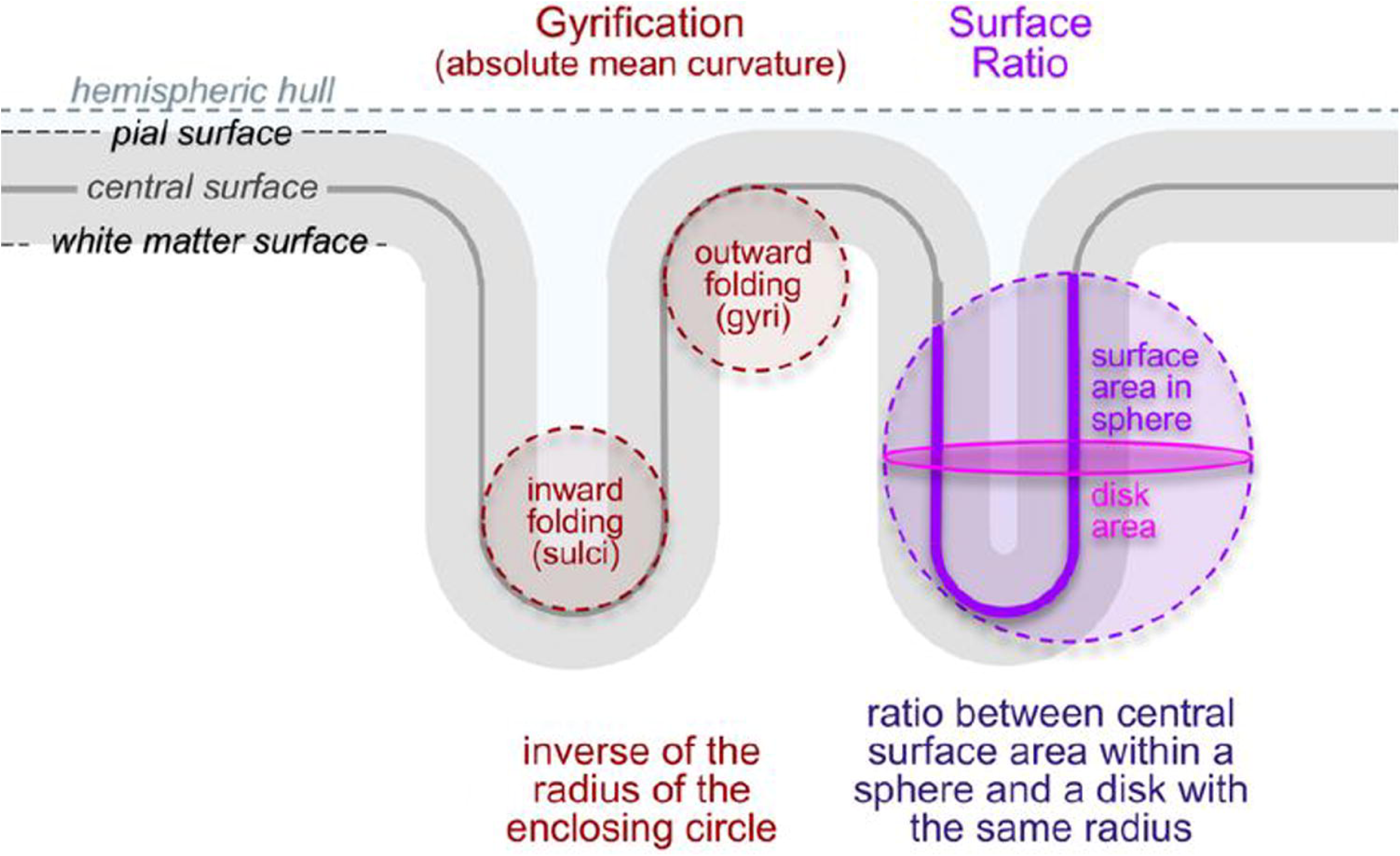
Graphical representation of the two cortical folding measures provided by CAT, surface ratio and absolute mean curvature. Adapted with permission from Gaser et al. (2022).

### Image processing quality control

To make sure that the imaging data analyzed was of good quality, we used the *Data quality* tools of the CAT software, as well as the reports generated after the segmentation procedure, to identify participants that might have been processed inadequately. Of the initial sample of 67 participants, we excluded 3 (4.5%) after the data quality check. We excluded an additional participant from the HC group who had their MRI acquired almost one year after the clinical visit, thereby resulting in the final sample of 63 participants.

### Clinical and antipsychotic medication data

We obtained the scores from the Positive and Negative Syndrome Scale (PANSS) (Kay, Fiszbein, and Opler 1987) that was administered to the patients at the clinical visit. We also gathered information about the patients’ daily antipsychotic medication intake, illness duration (years of difference between the date of symptom onset and the clinical visit),^1^ and tobacco and cannabis use.

### Statistical analysis

First, we conducted ANOVA and chi-squared analyses, where appropriate, to compare the sociodemographic and clinical data between groups. Second, we explored brain gyrification differences between the three groups (SCH-H, SCH-NH, and HC) by means of a whole-cortex ANCOVA SBM analysis of the SR and AMC maps, with age, sex, and education level as covariates. Third, to address potential associations between the PANSS scores and brain gyrification, we conducted whole-cortex multiple regression SBM analyses of the SR and AMC maps in the SCH-H and SCH-NH groups separately; in each one of these analyses, we employed one of the four PANSS global scores (for positive, negative, and general subscales, as well as total score) as the independent variable, and age, sex, and education level as covariates. To test the robustness of these analyses, we repeated all of those that yielded significant results with the addition of other potentially confounding variables, namely antipsychotic medication, illness duration, tobacco use, and cannabis use, one at a time. Finally, for all the items belonging to the subscales that reached significance in the previous analyses, we performed additional whole-cortex multiple regression SBM analyses, using the score for each item as the independent variable, and age, sex, and education level as covariates. In all the SBM analyses, we employed a vertex-level cluster-defining threshold of p < 0.001 and a family-wise error (FWE)-corrected cluster-level threshold of p < 0.05.

### Ethics statement

The study was approved by the local research ethics committee and complied with the provisions of the World Medical Association Declaration of Helsinki. All the participants and/or their legal representatives provided written informed consent.

## Results

### Sociodemographic and clinical data comparison between groups

A comparison of sociodemographic and clinical data between the HC, SCH-NH, and SCH-H groups is shown in Table 1. While we did not find significant differences between groups in age or sex distribution, SCH-H patients were significantly less educated than HC and smoked more tobacco than HC and SCH-NH. Additionally, SCH-H patients had a longer illness duration, higher antipsychotic medication intake, and higher PANSS scores than SCH-NH patients.

**Table 1.**
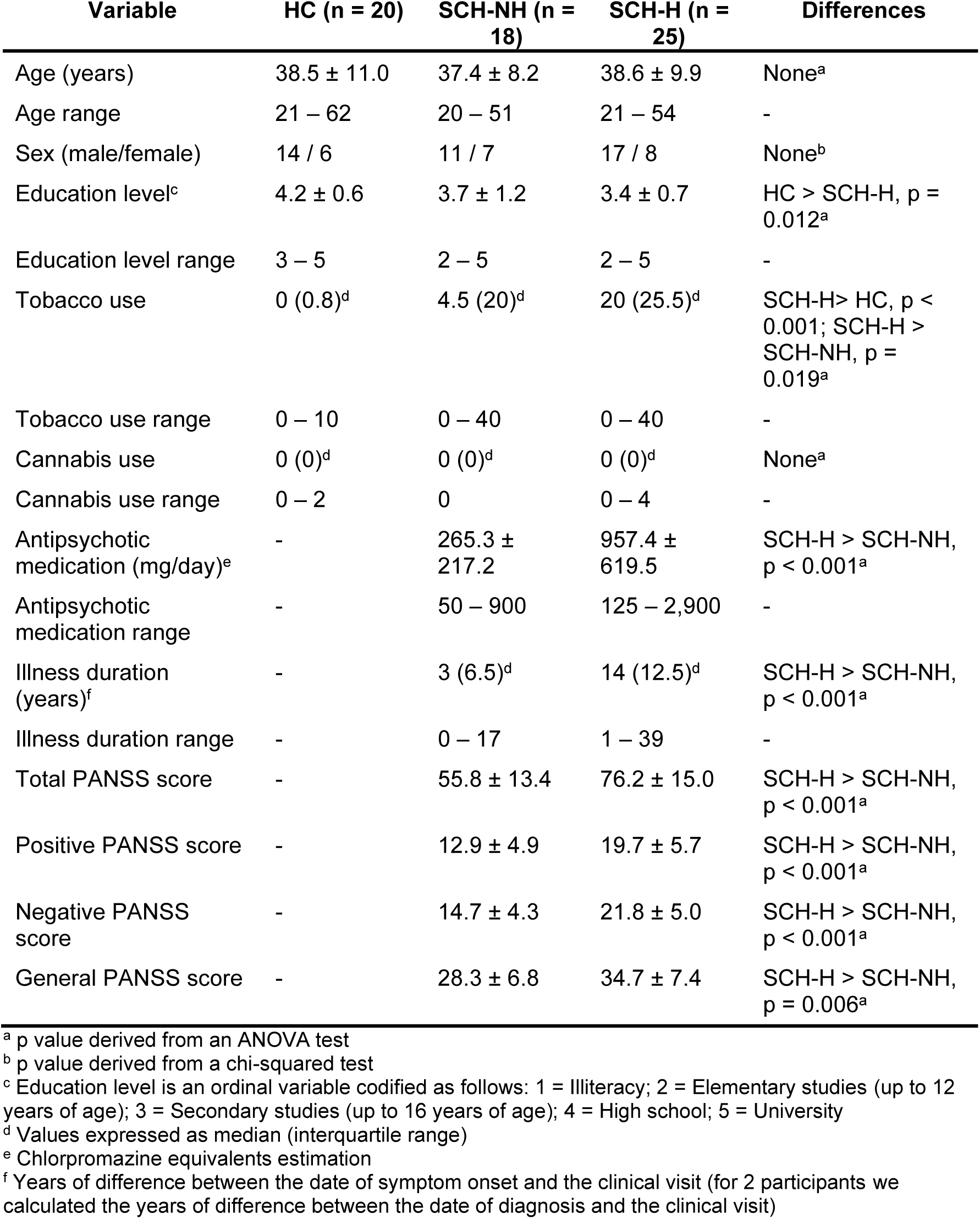
Sociodemographic and clinical data comparison between healthy controls (HC), schizophrenia patients without hallucinations (SCH-NH), and schizophrenia patients with treatment-resistant auditory verbal hallucinations (SCH-H). Values are expressed as mean ± standard deviation unless otherwise specified.

### Brain gyrification differences between groups

The analysis of brain gyrification differences between groups revealed that SCH-H patients displayed higher SR than SCH-NH patients and HC in the left orbitofrontal cortex. Moreover, SCH-H patients also had higher SR than HC in the right rostral middle and superior frontal regions. Conversely, both SCH-NH patients and HC showed higher SR than SCH-H patients in a cluster encompassing the right caudal middle frontal region and a more posterior part of the superior frontal, as well as precentral areas. Additionally, SCH-NH patients also had higher SR than SCH-H patients in another cluster that included right parietal and lateral occipital areas. No significant differences arose between SCH-NH patients and HC (Figure 2A and Table 2).

**Figure 2.**
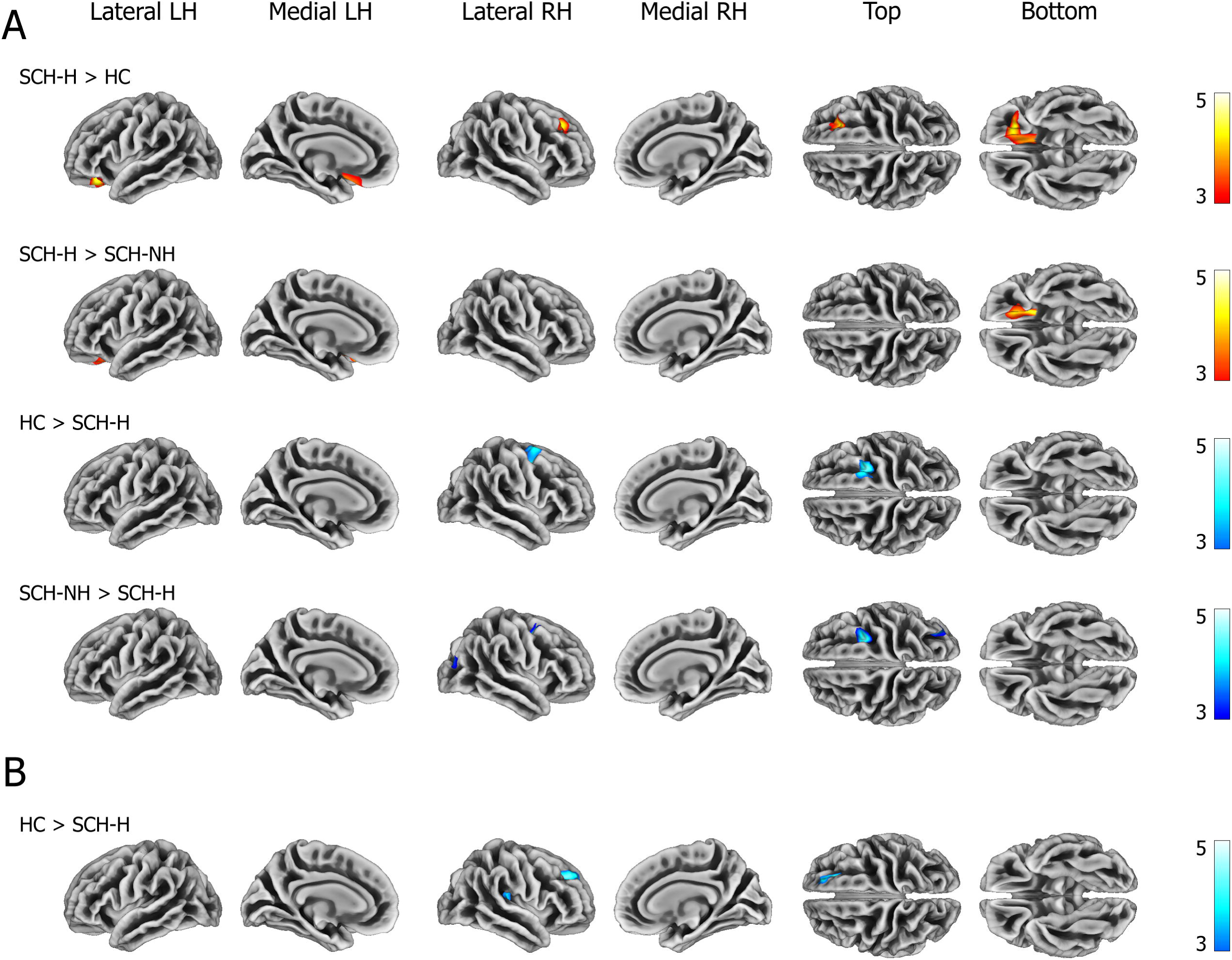
Brain gyrification differences between healthy controls (HC), patients without hallucinations (SCH-NH), and patients with treatment-resistant auditory verbal hallucinations (SCH-H), captured with **A)** surface ratio and **B)** absolute mean curvature. Red-yellow and blue colors depict regions where SCH-H patients showed higher and lower gyrification, respectively, than the other groups. We identified these regions by means of whole-cortex ANCOVA surface-based morphometry analysis, including age, sex, and education level as covariates. We employed a vertex-level cluster-defining threshold of p < 0.001 and a family-wise error (FWE)-corrected cluster-level threshold of p < 0.05. Color bars represent T values. See Table 2 for a list of the regions depicted here.

**Table 2.** List of regions showing significant brain gyrification differences between healthy controls (HC), patients without hallucinations (SCH-NH), and patients with treatment-resistant auditory verbal hallucinations (SCH-H), captured with either the surface ratio or the absolute mean curvature measures. See Figure 1 for further details and a graphical representation of these results.

| Cluster size (p value) | Peak T value | Region (cluster % within region) |
| --- | --- | --- |
| SCH-H > HC (surface ratio) |  |  |
| 1,894 (< 0.001) | 4.4 | Left lateral orbitofrontal (58%) |
|  |  | Left medial orbitofrontal (42%) |
| 1,383 (0.001) | 4.4 | Right rostral middle frontal (58%) |
|  |  | Right superior frontal (37%) |
|  |  | Right caudal middle frontal (6%) |
| SCH-H > SCH-NH (surface ratio) |  |  |
| 1,063 (0.005) | 4.3 | Left lateral orbitofrontal (67%) |
|  |  | Left medial orbitofrontal (33%) |
| HC > SCH-H (surface ratio) |  |  |
| 1,576 (< 0.001) | 4.2 | Right superior frontal (53%) |
|  |  | Right caudal middle frontal (27%) |
|  |  | Right precentral (19%) |
| SCH-NH > SCH-H (surface ratio) |  |  |
| 1,044 (0.006) | 4.8 | Right caudal middle frontal (61%) |
|  |  | Right superior frontal (25%) |
|  |  | Right precentral (14%) |
| 681 (0.040) | 4.2 | Right inferior parietal (86%) |
|  |  | Right lateral occipital (13%) |
|  |  | Right superior parietal (1%) |
| HC > SCH-H (absolute mean curvature) |  |  |
| 744 (0.019) | 4.2 | Right superior frontal (89%) |
|  |  | Right rostral middle frontal (11%) |
| 736 (0.020) | 3.7 | Right insula (52%) |
|  |  | Right transverse temporal (25%) |
|  |  | Right supramarginal (22%) |

As for the differences in AMC, we identified two clusters that showed more gyrification in HC than in SCH-H patients; both clusters were located in the right hemisphere, one of them encompassing the superior and rostral middle frontal regions and the other the insula, the transverse temporal gyrus, and the supramarginal gyrus (Figure 2B and Table 2). We did not find differences between SCH-H and SCH-NH patients or between SCH-NH and HC.

### Brain gyrification and global PANSS scores

In the SCH-H group, we found an association between higher SR in the right orbitofrontal region and lower total PANSS scores. This association appeared to be driven by the scores on the positive subscale, since higher SR in the right orbitofrontal region was also associated with a lower score on this subscale (Figure 3 and Table 3). We found these relationships to be robust, since they were still significant after the inclusion of additional potentially confounding variables, namely antipsychotic medication, illness duration, tobacco use, and cannabis use. No significant associations arose between SR and the negative or general subscales in the SCH-H group. Furthermore, we did not find PANSS scores to be significantly associated with SR in the SCH-NH group or with AMC in any group.

**Figure 3.**
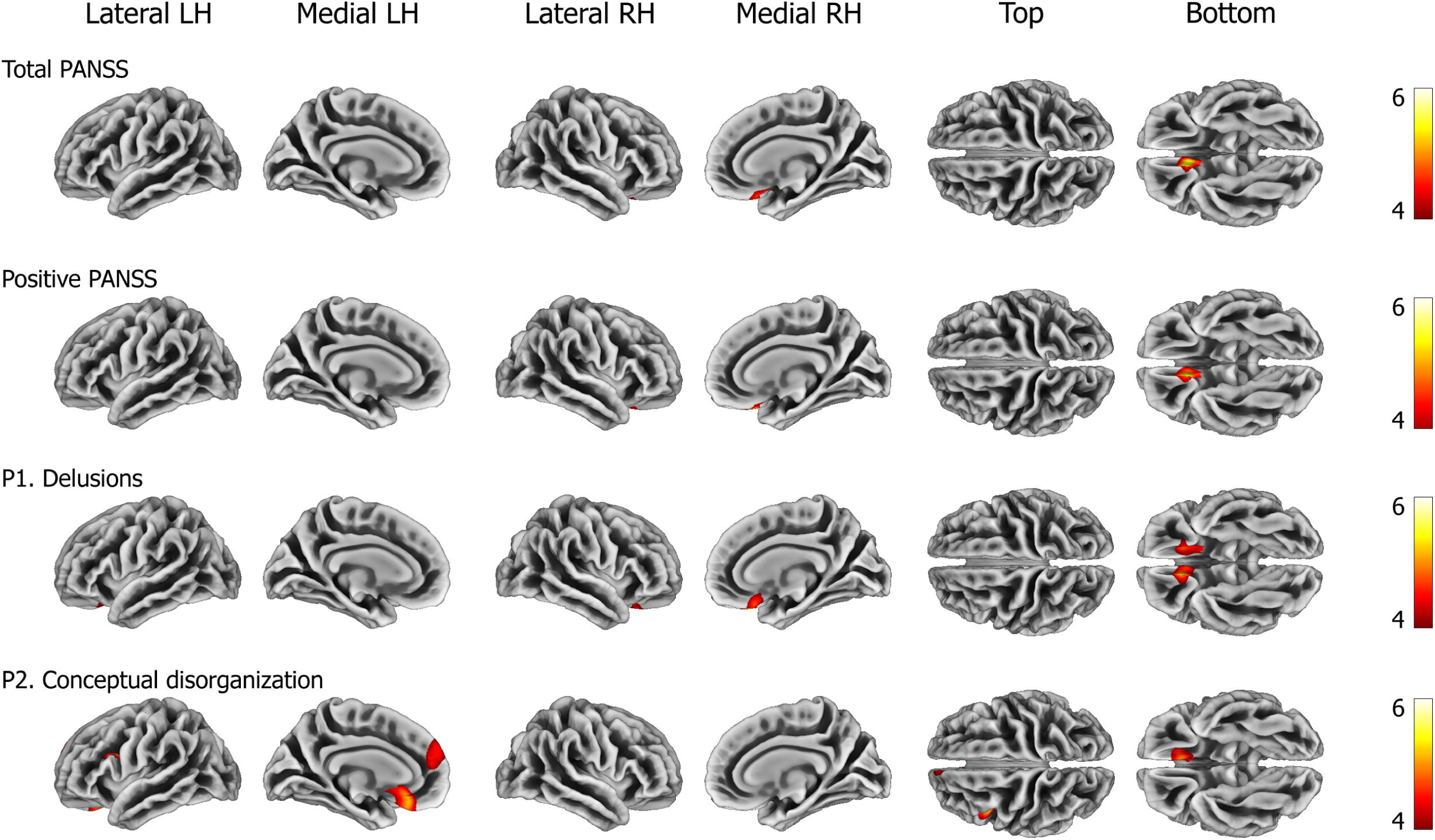
Associations between brain gyrification, as captured with the surface ratio measure, and scores on the Positive and Negative Syndrome Scale (PANSS) in patients with treatment-resistant auditory verbal hallucinations (SCH-H). In all cases, higher gyrification was associated with lower scores. We identified these regions by means of whole-cortex multiple regression surface-based morphometry analysis, including age, sex, and education level as covariates. We employed a vertex-level cluster-defining threshold of p < 0.001 and a family-wise error (FWE)-corrected cluster-level threshold of p < 0.05. Color bars represent T values. See Table 3 for a list of the regions depicted here.

**Table 3.** List of regions showing significant associations between brain gyrification (captured by the surface ratio measure) and several scores on the Positive and Negative Syndrome Scale (PANSS) in patients with treatment-resistant auditory verbal hallucinations (SCH-H). See Figure 2 for further details and a graphical representation of these results.

| Cluster size (p value) | Peak T value | Region (cluster % within region) |
| --- | --- | --- |
| **Total PANSS** |  |  |
| 953 (0.006) | 5.5 | Right medial orbitofrontal (58%)<br>Right lateral orbitofrontal (42%) |
| **Positive PANSS** |  |  |
| 994 (0.005) | 5.2 | Right lateral orbitofrontal (51%)<br>Right medial orbitofrontal (49%) |
| **Delusions (P1 item)** |  |  |
| 1,103 (0.002) | 5.5 | Right medial orbitofrontal (55%)<br>Right lateral orbitofrontal (45%) |
| 734 (0.020) | 4.5 | Left lateral orbitofrontal (50%)<br>Left medial orbitofrontal (50%) |
| **Conceptual disorganization (P2 item)** |  |  |
| 1,236 (0.001) | 5.0 | Left medial orbitofrontal (78%)<br>Left rostral anterior cingulate (19%)<br>Left lateral orbitofrontal (3%) |
| 690 (0.027) | 4.1 | Left superior frontal (100%) |
| 592 (0.049) | 5.5 | Left pars opercularis (90%)<br>Left rostral middle frontal (7%)<br>Left caudal middle frontal (3%) |

### SR and positive subscale items

Since the relationship between the positive subscale score of the PANSS and SR in the SCH-H group was significant and robust to the addition of several confounding variables, we decided to further explore potential associations between SR and the individual items belonging to the positive subscale.^2^ We found a significant association between higher SR at the bilateral orbitofrontal cortex and lower scores on the delusions item. Similarly, lower scores on the conceptual disorganization item were associated with higher SR in various clusters, with the largest one including the left orbitofrontal cortex—mainly its medial aspect—and the anterior cingulate cortex (Figure 3 and Table 3). No further significant associations arose with the rest of the items on the positive subscale, including the hallucinations item.

## Discussion

Here we report several brain gyrification differences between SCH-H patients and the SCH-NH and HC groups, mainly located at the frontal lobe. We found no differences between SCH-NH patients and HC. The region showing the most prominent gyrification differences was the orbitofrontal cortex, which seems to be hypergyrified in SCH-H patients. We additionally found, only within the SCH-H group, that a higher gyrification of the orbitofrontal cortex was associated with lower positive symptoms as measured with the PANSS, particularly with lower delusions and conceptual disorganization. Importantly, we identified most of the brain gyrification differences and associations with SR, a gyrification measure devised by Toro et al. (2008) and which, to our knowledge, was used here for the first time in a sample of patients with schizophrenia. While the AMC approach (Luders et al. 2006) allowed us to identify some gyrification differences between the SCH-H and HC groups, mainly located in the frontal lobe and the insula, it did not detect folding differences in the orbitofrontal cortex or between the two groups of patients. Moreover, the AMC approach yielded no significant associations with the PANSS scores.

The orbitofrontal cortex has been associated with schizophrenia multiple times; in fact, several studies have reported altered orbitofrontal gyrification patterns in psychosis (Nakamura et al. 2007; Bartholomeusz et al. 2013; Lavoie et al. 2014; Isomura et al. 2017). Our results with the SR measure are in agreement with other studies showing that the orbitofrontal cortex is more gyrified in patients with schizophrenia (Nenadic et al. 2015; Spalthoff, Gaser, and Nenadić 2018) and first-episode schizophrenia (Sasabayashi et al. 2017). However, it is worth noting that we found the hypergyrification of the orbitofrontal cortex to be specific to schizophrenia patients with treatment-resistant auditory verbal hallucinations. Although not in schizophrenia, a similar result was reported by Kubera et al. (2023), who observed hypergyrification of the left lateral orbitofrontal cortex in borderline personality disorder patients with auditory verbal hallucinations as compared with non-hallucinating patients. Ren et al. (2022) found lower cortical thickness in the bilateral lateral orbitofrontal cortices in schizophrenia patients with persistent auditory verbal hallucinations, which is somewhat consistent with our findings, considering that gyrification and cortical thickness tend to have an opposing relationship (Hogstrom et al. 2013; Gautam et al. 2015). The precise functions of the orbitofrontal cortex are still debated (Stalnaker, Cooch, and Schoenbaum 2015; Rudebeck and Rich 2018); however, it has been suggested as being implicated in decision-making (Padoa-Schioppa and Conen 2017; Noonan et al. 2017; Rudebeck and Rich 2018; Rolls 2023) and in socioemotional behaviors (Watson and Platt 2012; Rudebeck and Rich 2018; Rolls 2023). The orbitofrontal cortex is strongly interconnected with several parts of the brain, including the inferior frontal gyrus, the insula, and the anterior cingulate cortex (Rolls et al. 2022).

Even though the cerebral mechanisms that give rise to auditory verbal hallucinations are unclear (Gill et al. 2022), functional neuroimaging studies point primarily to language and auditory areas, such as the inferior frontal gyrus and the superior temporal gyrus (Jardri et al. 2011; Richards et al. 2021). Yet some studies have shown that the orbitofrontal cortex is also active during auditory verbal hallucinations (Silbersweig et al. 1995; Parellada et al. 2008; Hugdahl et al. 2023), and some case studies have reported that patients with orbitofrontal cortex seizures (La Vega-Talbot, Duchowny, and Jayakar 2006) or lesions (Shah, Manning, and Sharma 2015) may also manifest auditory hallucinations. Since auditory verbal hallucinations have also been associated with enhanced activity of the insular cortex (Jardri et al. 2011; Barber, Reniers, and Upthegrove 2021), some authors have suggested that a malfunctioning salience network may be related to the hallucinatory experience (Palaniyappan and Liddle 2012; Barber, Reniers, and Upthegrove 2021). Of note, the salience network is mainly formed by the insula and the anterior cingulate cortex (Barber, Reniers, and Upthegrove 2021). Even if we do not consider the orbitofrontal cortex to be at the core of auditory verbal hallucinations, there seems to be a good deal of evidence suggesting a role for this region in hallucinatory experiences. Our gyrification result provides additional evidence and may indicate that SCH-H patients present connectivity alterations in that region. Indeed, connectivity abnormalities in the frontal lobe have been previously reported in treatment-resistant schizophrenia patients (Alonso-Solís et al. 2015; Tuovinen and Hofer 2023). If we assume that cortical folding is proportional to neuronal connectivity, at least locally (Van Essen 1997; Herculano-Houzel et al. 2010; Dauvermann et al. 2012), it is possible that SCH-H patients have an excess of connectivity within the orbitofrontal cortex as compared with SCH-NH patients and HC. Therefore, and given the strong interconnections that the orbitofrontal cortex has with auditory and language areas—such as the inferior frontal and superior temporal gyri—and with the regions that form the salience network (Rolls et al. 2022), it may be hypothesized that an aberrant and/or excessive wiring of the orbitofrontal cortex leads to incorrect interpretation of internal stimuli (Menon 2011), which may give rise or contribute to the auditory verbal hallucinations experienced by the SCH-H patients. An alternative hypothesis is that our results reflect the effect of a compensatory mechanism by which the brains of SCH-H patients may attempt to restore lost connections—or at least preserve the existing ones—, perhaps in response to atrophy in nearby areas. In this way, the natural process of age-related gyrification loss (Hogstrom et al. 2013) would be softened in order to maximize the available space required for those connections.

Greater gyrification of the orbitofrontal cortex, as measured with SR and only within the SCH-H group, was also associated with less severe delusions and conceptual disorganization, further highlighting the importance of considering the folding of this region in schizophrenia. In particular, less severe delusions were associated with greater gyrification of the bilateral orbitofrontal cortex in both its lateral and medial aspects. A similar result, localized at the right lateral orbitofrontal cortex, was observed by Hua et al. (2022) in a sample of participants at risk of developing psychosis. Lesions to the left orbitofrontal cortex have also been associated with delusional misidentifications (Tanabe et al. 2018). Regarding conceptual disorganization, which may be defined as difficulties in directing thoughts toward a goal (Kay, Fiszbein, and Opler 1987), it was mainly associated with a cluster involving the left medial orbitofrontal cortex and the rostral anterior cingulate cortex. A previous study reported reduced white matter integrity of the cingulum tract in a group of first-episode psychosis patients with conceptual disorganization symptoms (Pan et al. 2021). As mentioned above, the anterior cingulate cortex is part of the salience network (Barber, Reniers, and Upthegrove 2021) and is interconnected with the medial orbitofrontal cortex, forming a pathway that seems to be particularly important for goal-directed actions (Rolls 2023). Thus, this cortical folding relationship with delusions and conceptual disorganization in SCH-H patients may also reflect underlying connectivity issues in the orbitofrontal cortex. In the case of conceptual disorganization, this may specifically involve the medial orbitofrontal cortex and the anterior cingulate cortex. However, it is somewhat striking that we found the severity of positive symptoms to be inversely correlated with orbitofrontal cortex folding in the SCH-H group, as this may seem to contradict our previous result, by which the higher folding of the orbitofrontal cortex appeared to be indicative of a more severe disease. One possibility is that hypergyrification of the orbitofrontal cortex represents a more severe pathology when considering a broader spectrum of the disease (i.e., taking into account SCH-NH patients), but that this relationship does not stand within the SCH-H group, in which a more folded orbitofrontal cortex would actually be associated with less severe delusions and conceptual disorganization symptoms. In fact, we did not find any association between brain folding and the severity of hallucinations in SCH-H patients, although it is important to note that the PANSS assesses not only auditory hallucinations but also other hallucinatory modalities. Overall, this explanation might fit better with the compensatory hypothesis mentioned previously, suggesting that patients who achieve higher compensation rates also display less severe positive symptoms than other patients with treatment-resistant auditory verbal hallucinations.

The methodology followed in this study also allowed us to accurately compare two different approaches to quantifying cortical folding, namely the SR and AMC measures. We applied these two measures to the same sample, using the same software, and after the same initial image processing, which allowed us to reliably compare them. Importantly, we obtained the most relevant results with the SR measure, which seemed to capture associations that would have otherwise gone unnoticed with the AMC measure. While we detected some gyrification differences between SCH-H and HC with the AMC approach, no differences arose between the two groups of patients or within groups in association with symptoms. We expect that future investigations will employ the promising SR approach in larger schizophrenia samples to evaluate the robustness of the results presented here and potentially reveal new brain regions of interest. Future studies may also help to provide an answer to the questions raised throughout this section.

This study has several strengths. First, we analyzed a well-characterized schizophrenia sample that included both patients with treatment-resistant auditory verbal hallucinations and patients without hallucinations; second, we employed a whole-cortex approach that allowed us to locate gyrification differences and associations in a precise and unbiased manner; third, we controlled the potentially confounding effects of several variables, namely antipsychotic medication, illness duration, tobacco use, and cannabis use; and finally, we acquired the MRI of all the participants in the same scanner and, in almost all cases, on the same day as the clinical visit, thereby maximizing the homogeneity of the sample. The main limitation of this study is the relatively small sample that we employed. Moreover, since we did not analyze longitudinal data, we were not able to evaluate potential cortical folding changes over time.

Overall, our findings confirm and extend the results of previous studies by showing that the folding of the orbitofrontal cortex is an important brain structure biomarker in schizophrenia. The SR measure to estimate brain gyrification seems to be more sensitive than alternative measures and shows great promise in the study of psychotic disorders.

## Data Availability

All data produced in the present study are available upon reasonable request to the authors.

## Footnotes

1 For two participants (one SCH-H and one SCH-NH), we calculated the years of difference between the date of diagnosis and the clinical visit, since we did not know the date of symptom onset.

2 Since we did not have the individual item scores for one participant, we carried out these analyses with 24 SCH-H patients.

